# Bacteriological quality of drinking water and associated factors in Debre Markos town, North West, Ethiopia

**DOI:** 10.1101/2025.03.24.25324515

**Authors:** Bantegzie Selabat, Hylemariam Mihretie, Brhanu Kassanew, Abeba Mengist

## Abstract

**Background:** Drinking of biologically deteriorated water is the major cause of water-borne disease globally.

**Objective:** The aim of this study was to assess the bacteriological quality and associated factors of drinking water in Debre Markos town, Northwest, Ethiopia.

**Materials and Methods:** A cross-sectional study design was conducted from June 2021 to April, 2022. Purposive, simple random and stratified sampling techniques were used to collect a total of 211 water samples from water sources and households. After about 400ml of water sample were collected, water samples were analyzed by membrane filtration method using membrane lauryl sulphate broth media at Debre Markos water supply and sewerage service laboratory to identify total coliform and thermotolerant coliform. Data was collected and double entered to Epi data version 3.1 and exported to SPSS version 25 software for statistical analysis. Logistic regression with 95% CI was used to show the statistical association. P value <0.05 was considered as statistically significant.

**Result:** This study result indicates that 84/211 (39.8%) and 74/211 (35.5%) of water samples were positive for total coliform and thermotolerant coliform, respectively. Multivariable logistic regression revealed that household samples from hand pump beneficiaries (AOR=11; 95% CI: (3.012, 40.368)) (p<0.001) and stored in not covered jerry can (AOR=25.6; 95% CI: (3.322, 37.312)) (p<0.002) were more likely to be contaminated with coliform. In addition, water samples from not covered storage containers (AOR=3; 95% CI: (1.015, 9.156)) (p<0.047) and water samples from households those not used water treatment in home (AOR=16.9; 95% CI: (2.754, 24.804)) (p<0.002) were more likely to be contaminated with coliforms.

**Conclusion:** This study revealed that bacteriological quality of drinking water deteriorates from sources to point of use. The presence of thermotolerant coliform in the drinking water samples demonstrates the presence of pathogenic microorganisms that would be a threat to individual consuming the water. Thus, regular monitoring of bacteriological quality of drinking water sources and health education programs on water quality should be enhanced to improve the quality of drinking water.

## INTRODUCTION

Microbiological quality refers to the presence of organisms that cannot be seen by the naked eye, such as bacteria, viruses and protozoa. Drinking water does not cause an infectious disease if it is free from indicator microorganisms [1]. Water is highly vulnerable to pathogens as the water travels a long distance in the water cycle, and it can be contaminated easily in any one of the many points in the way from sources to point of use [2].

Approximately 1.8 billion people use fecal contaminated water sources globally, with the majority living in low and middle income countries [3]. Contamination of water resources by pathogenic bacteria causes more than 80% of the human diseases in the world, and it affects individuals, households, communities and countries [4].

About 60-80% of the population of Ethiopia suffers from serious water-related diseases [5] and above 75 % of the health problems are due to communicable diseases attributed to unsafe water supply unhygienic waste management, particularly human excreta [6]. Ministry of health estimated that 6000 children die each day from diarrhea and dehydration [7].

The health sector of Debre Markos town frequently reports that water related diseases such as Diarrhea and typhoid fever and these are among top-ten leading cases in town. The reports of these diseases are indicators that the population of the town is suffering from waterborne diseases, this is probably due to poor drinking water quality (Debre Markos compressive specialized hospital report, 2021). Although the problem of bacteriological quality of drinking water quality exits, to date, there is paucity of published study conducted in the study area regarding the bacteriological quality and associated factors of drinking water. Therefore, this study was conducted to assess the bacteriological quality and associated factors of drinking water in Debre Markos town.

## MATERIALS AND METHODS

### Study design, period and setting

A cross-sectional study was conducted to assess bacteriological quality of drinking water in Debre Markos town, North West Ethiopia from June, 2021 to April, 2022.

The main water supply of the town is groundwater distributed through pipe. The residents of the town are currently getting drinking water supply service from Sentera and Wutrin water supply network. In the distribution system, water enters into reservoirs located at three places and there are eleven pressure zones in the town. In additions, hand-pumps, springs and public taps (common taps) are public sources for drinking for the community of Debre Markos town.

### Inclusion and Exclusion criteria

All public drinking water sources managed by Debre Markos water supply and sewerage service and were found giving service for the community and all households whose owner’s age were more than 18 years and were volunteered to give consent included in the study. Water sources not functional at the time of sample collection and broken water container were excluded from the study.

### Sample size determination and sampling techniques

#### Sample size determination

The samples size for this study was determined using single population proportion formula for household water samples:

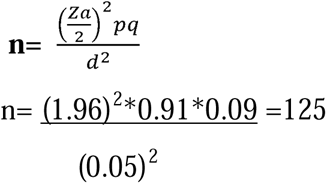

By assuming z = 95% confidence interval i.e.1.96, p = previous proportion of contaminated water i.e. 0.91 [8], q= 1-p i.e. 0.09, d = margin of error (0.05), n= household containers (125), and purposively include household tap waters (61) and other water sources (25), the total sample size was 211. The sample size of water for household containers were proportionally allocated to households those benefited drinking water from public water sources and households those benefited drinking water from household tap to keep representativeness.

### Sampling techniques

Purposive and simple random sampling techniques were used for sources and households, respectively. Stratified random sampling technique was also used to recruit the study households those benefited drinking water from public water sources such as springs (n=4), hand pumps (n=10) and public taps (n=2). And households those benefited drinking water from household tap were selected by lottery method to keep the representativeness. In this way, 5 pressure zones (45%) such as 2, 3, 4, 5 and 6 were selected from the total 11 pressure zones. Then the sample size was allocated proportionally to each pressure zone depending on the total number of households, and water samples were taken from both taps and containers.

Sampling frame was obtained from Debre Markos water supply and sewerage service registration books. Water samples were collected three times from each sampling point. Overall, 211 water samples (125 from households’ container, 61 from household taps and 25 from sources) were collected in the study.

#### Data Collection tool

Data was collected by trained medical laboratory technologist. Data were collected from sources by using prepared checklist and data from households were collected by using a structured questioner. For household’s data, after the respondents agreed to take part in the study and signed on informed written consent form, sociodemographic characteristics and household sanitation and water handling practices data were collected. Questionnaires were first developed in English and translated into Amharic (local language), and then the responses were translated back into English language.

#### Sample collection techniques

In the study area, water samples were collected from all public water sources serving the population of study area and from households. The method of sample collection from each sampling source was according to WHO guidelines for drinking water quality assessment [9].

About 400ml water samples were collected in sterilized glass bottles that was washed and rinsed thoroughly with nitric acid and distilled water. The chlorinated water sample was collected using glass bottles sterilized after adding 0.1 ml sodium thiosulfate. Sample collection techniques were varied according to the type of water sample.

#### Household tap/public tap

The tap outlet was cleaned using a cloth and allow for maximum flow for two minutes; and then Outlet was sterilized with flame using a lighter. The tap again was allowed to flow for 1-2 minutes at medium flow rate. Using sterile glass bottle water was collected and delivered to laboratory within one hour.

#### Spring

Samples from springs was taken according to surface water sampling procedure by dipping the sampling container to 20 cm of the water body. In each round of sampling of springs, one sample will be taken at the center and the other two samples from the left and right edges [10].

#### Hand pump

Before taking samples from hand pump, first, the mouth of hand pump was cleaned using a cloth and allow for maximum flow for two minutes; and then the mouth of the hand pump was sterilized with a flame from ignited alcohol-soaked cotton. The water again was allowed to flow for 1-2 minutes at medium flow rate. Using sterile glass bottle water was collected and delivered to laboratory [11].

#### Borehole

The hydrant valve had opened and bibcock until water runs. Bibcock was shut and allowed at least 1 minute and then, opened fully to allow water to flow at least 2 minutes. About 400ml water was collected aseptically.

#### Reservoir Water

A sterile bottle was tied on to a rope that was tied with a sterilized stone or a sterilized heavy piece of metal at its lower tip. The stone or heavy piece of metal is to provide weight for the rope to get into the water. The cap of the bottle was removed and lowered the bottle into the well to a depth of about 1meter and the bottle was raised out and carefully replaced the cap.

#### Household storage containers

The water-drawing cup was wiped using a clean cotton pad and sterilized with a flame from ignited alcohol-soaked cotton. About 400ml of water sample was taken using sterilized glass bottle. All water samples collected by trained laboratory technologists.

#### Bacteriological water analysis and procedures

After water samples were taken from sources and households, bacteriological quality of the water were analyzed according to the standard methods described by the American Public Health Association guideline (APHA). Water samples were analyzed for total coliform and thermotolerant coliform using the membrane filter technique as outlined by the American Public Health Association guideline [12].

All collected samples from water sources and households were transported to Debre Markos water supply and sewerage service laboratory within one hour of collection using a cold box with ice packs for storage. Samples were examined within 6 hours of collection according to standard examination methods to avoid the growth or death of coliforms in the samples [13]. Each water sample was mixed thoroughly by shaking for 30 minutes. A hundred ml of water sample was filtered through 0.45 μm a membrane filter in a vacuum filtration apparatus for each of total coliform and thermotolerant coliform, and all the filters were transferred to the membrane lauryl sulphate broth media. Then, plates for TC and TTC counts were incubated at 37 °C and 44 °C, respectively, for 24 hrs. Up on completion of the incubation period, yellow color colonies on the surface of the filter paper were counted both for TC and TTC [14].

#### Data quality control

All the questions in a structured questionnaire were prepared in a clear and precise way and translated into the local language (Amharic). Training was given for data collector how to collect data from study participants. Supervision was conducted on data collector and the assessment of water handling practices of the households of the community was collected using a structured questionnaires. Before sample collection, sampling bottles was properly labeled with households ID, source of water and date and hour of collection for easily identification of the sample.

Sample bottles was rinsed with tap water, thereafter soaked in 10% HNO3 for 24 h, and finally rinsed with de-ionized water. Sample transportation was also performed with cold box to maintain the quality. To keep the quality of media any physical changes (growth of microorganism) on culture media was assessed, sterility of media was checked through overnight incubation and expiration date of media was checked. Standard operation producers were strictly followed during reagent preparation, incubation and recording result. To keep the validity of the analysis, distilled water was included as control at the same time during the analysis.

#### Data Processing and Analysis

Bacteriological quality of drinking water in Debre Markos town were analyzed and compared with Ethiopian standard and WHO guideline. Drinking water (potable water) should free from coliform per 100ml water sample [9, 15]. The data were entered into Epi data version 3.1 and exported to SPSS version 25 software for statistical analysis. The results were presented by using tables. Frequencies and percentages were calculated using descriptive statistics to summarize results of categorical variables. Logistic regression was used to show statistical association between the dependent and independent variables. Crud odds ratio (COR) was analyzed by binary logistic regression and those Variables with their p-value <0.25 were candidates for multivariate logistic regression. Then multivariate logistic regression was used in terms of adjusted odds ratio (AOR) with 95% confidence intervals to minimize the confounding effect and p-value < 0.05 was considered as statistically significant.

## RESULTS

### Bacteriological quality of drinking water sources and households’ drinking water

In this study, the total water samples to analysis the bacteriological quality of drinking water were 211 and 84/211 (39.8%) of the drinking water sample were positive for TC and 75/211 (35.5%) of water sample were positive for TTC. The prevalence of TTC were 6/25 (24%), 17/61(28%), and 52/125(42%) from sources, household taps and households’ container respectively and did not compliance with the WHO guidelines [9].

According to risk classification for thermotolerant coliforms [16], mean value of thermotolerant coliforms in hand-pump were in high risk categories and the mean value of thermotolerant coliforms of household taps and households’ containers water samples were in the low-risk categories. Higher mean number of TC and TTC counts were observed particularly in unprotected springs and protected hand pumps. Neither total coliform nor fecal coliforms were detected from samples taken from the reservoirs and public taps. This finding showed that the bacteriological deterioration of water were increased from the main sources to household taps, and from household taps to point of use (Table 1).

**Table 1:** Prevalence and mean number of TC and TTC colonies/100ml of drinking water samples in Debre Markos town, North West, Ethiopia.

| Prevalence and mean number of TC and TTC/100ml water sample |  |  |  |  |  |  |
| --- | --- | --- | --- | --- | --- | --- |
| Type of source | N | TC [n (%)] | Mean number<br>of TC/100ml | TTC [n (%)] | Mean number<br>of TTC/100ml | Risk<br>Level |
| Boreholes | 6 | 5(83.3%) | 95.33 | 0(0%) | 0.00 | NR |
| Reservoirs | 3 | 0(0%) | 0.00 | 0(0%) | 0.00 | NR |
| Protected springs | 2 | 1(50%) | 45 | 1(50%) | 6 | LR |
| Unprotected springs | 2 | 2(100%) | 145 | 2(100%) | 7.5 | LR |
| Hand-pump | 10 | 5(50%) | 100.1 | 3(30%) | 24 | HR |
| Public tap | 2 | 0(0%) | 0.00 | 0(0%) | 0.00 | NR |
| Taps at each HH | 61 | 17(28%) | 7.77 | 17(28%) | 3.59 | LR |
| HH containers | 125 | 54(43.2%) | 11.9 | 52(42%) | 6.97 | LR |

Note. N=number of water samples, TC= total coliform count, TTC= thermotolerant coliforms count, NR= no-risk, LR= low-risk, and HR= high-risk

### Hygienic practices related to water collection and transportation

Participant were benefited drinking water from household taps, hand-pumps, springs and public tap; 61(48.8), 40(32%), 16(12.8%) and 8(6.4%) of them were water sample collected respectively. The most commonly preferred type of water collection container in study area was jerry can 98(78.4%). Out of total 125 respondents, 87(69.6%) of the respondents cleaned their containers before collection and 53(42.4%) of the respondents collected once a day (Table 2).

**Table 2:**
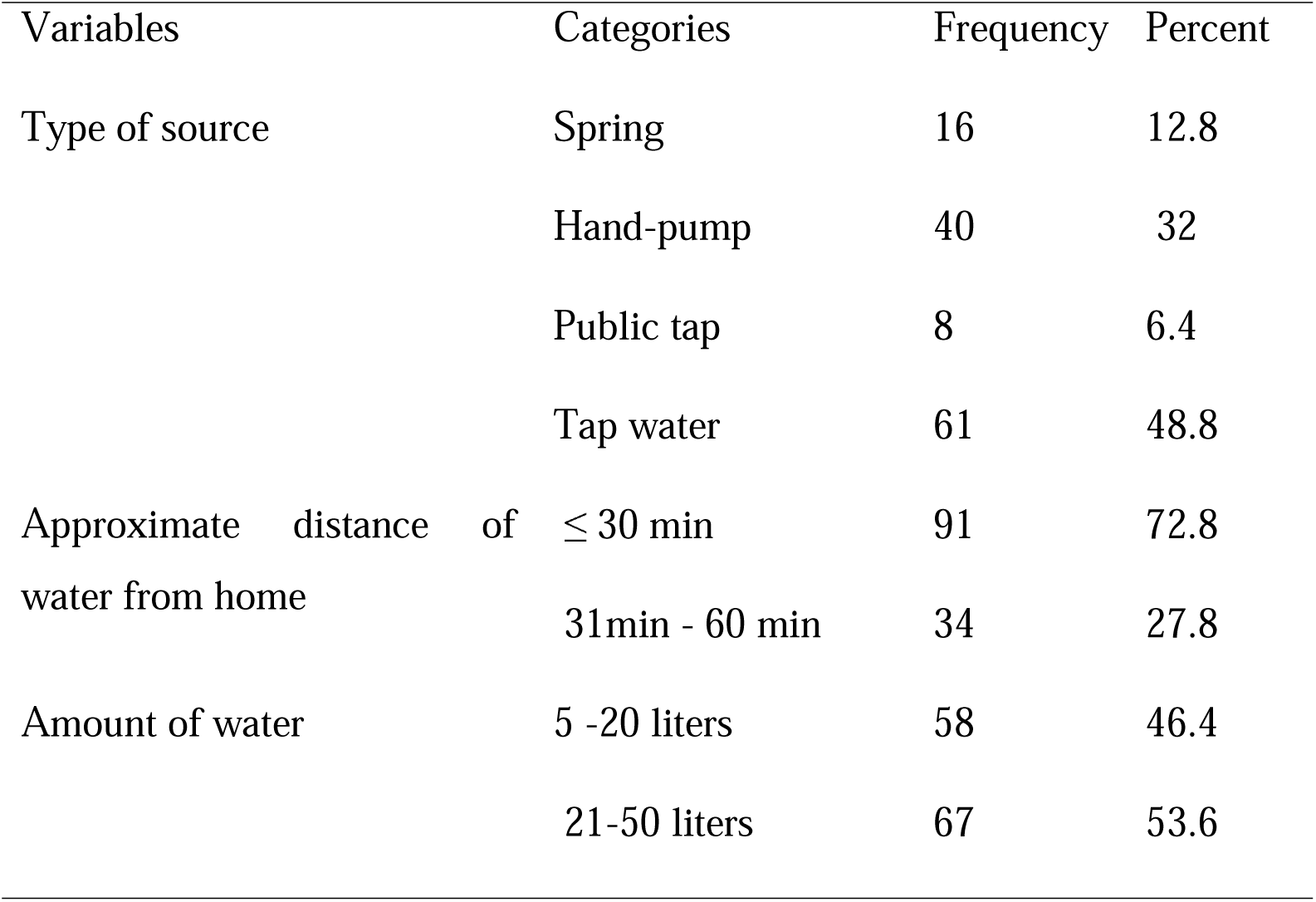
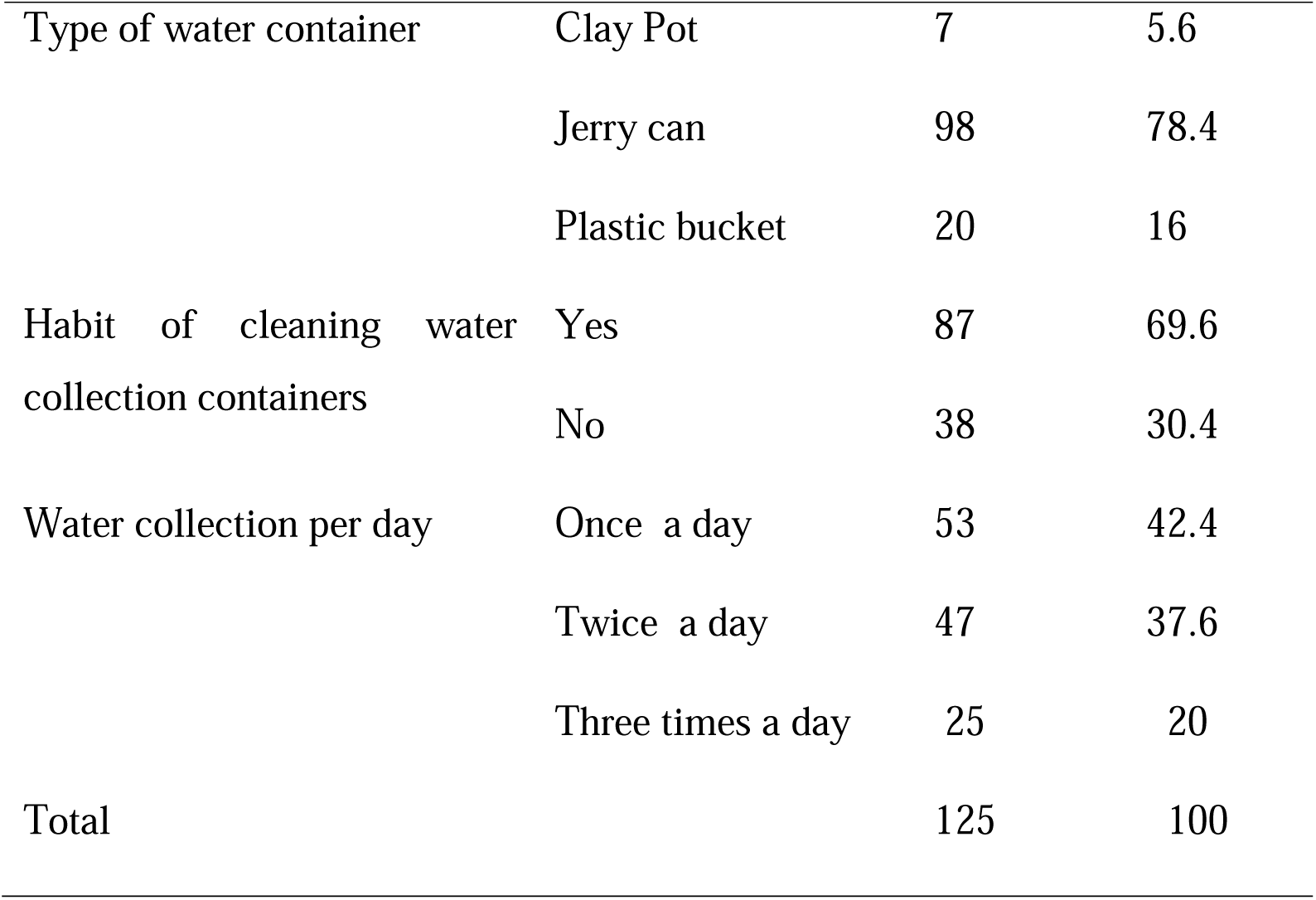
Water handling practices in related to collection and transportation in Debre Markos town, North West, Ethiopia.

### Hygienic practices related to drinking water storage and usage

Majority of respondents were used separate containers 84(67.2%) for drinking water from other intended purpose water. Respondents used for drinking water storage in household jerry can 91(72.8%), clay pot 16(12.8%) and plastic bucket 18(14.4). Participants used method of dipping and pouring from storage containers 52(41.6%) and 73 (58.4%) respectively. Concerning the placement of drinking cup 85(68%) put on the table, 15 (12%) placed on the floor and 25(20%) hung on the wall. Majority of respondents of drinking water were not treated in home 101 (80.8%) (Table 3).

**Table 3:** Water handling practices related to storage and usage in Debre Markos town, Ethiopia.

| Variables | Categories | Frequency | Percent |
| --- | --- | --- | --- |
| Use soap/ashes after toilet | Yes | 88 | 70.4 |
|  | No | 37 | 29.6 |
| Separate container for drinking | Yes | 84 | 67.2 |
|  | No | 41 | 32.8 |
| Type of water storage | Clay Pot | 16 | 12.8 |
|  | Jerry can | 91 | 72.8 |
|  | Plastic bucket | 18 | 14.4 |
| Cleaning of storage | Rarely | 14 | 11.2 |
|  | Sometimes | 67 | 53.6 |
|  | Always | 44 | 35.2 |
|  | Pouring | 73 | 58.4 |
| Water drawing | Dipping | 52 | 41.6 |
|  | Yes | 78 | 62.4 |
| Storage cover | No | 47 | 37.6 |
|  | On the table | 85 | 68 |
| Placement of drinking cup | On the floor | 15 | 12 |
|  | Hung on the wall | 25 | 20 |
|  | Less a day | 26 | 20.8 |
| Days water store | For a day | 50 | 40 |
|  | More than a day | 49 | 39.2 |
| Water treatment in home | Yes | 24 | 19.2 |
|  | No | 101 | 80.8 |
| Total |  | 125 | 100 |

### Bacteriological quality of drinking water with socio-demographic characteristics

In this study, the highest respondents constituted the age group 31-45 years. In age group > 45 years respondents, water samples were relatively more contaminated with indicator bacteria than the age group of 18-30 and 31-45years. Of total participants, 75.2% of the households head were male and 70.4% of the households head marital status were married. The educational status of the household heads, 48% of households heads attained college and above, 19.2% of households heads attained secondary education, and 16.8% of household heads attained primary education, while 16% of household heads were illiterate. Bacteriological deterioration of drinking water were high in illiterate heads of households than educated household heads. Above half of the participant households were staying with 1-3 family size and 28% and 20.8% were staying with 4-5 and > 5 family size respectively (Table 4).

**Table 4:** Bacteriological quality of drinking water with socio-demographic characteristics in Debre Markos town, Ethiopia.

| Variables | Categories | Total |  | Thermotolerant |  |  |
| --- | --- | --- | --- | --- | --- | --- |
|  |  | coliform count/100ml |  | coliform count/100ml |  | Total |
|  |  | Yes | No | Yes | No |  |
| Age of the respondent | 18-30 | 22(46.8%) | 25(53.2%) | 21(44.7%) | 26(55.3%) | 47 |
|  | 31-45 | 16(33.3%) | 32(66.7%) | 16(33.3%) | 32(66.7%) | 48 |
|  | >45 | 16(53.3%) | 14(46.7%) | 15(50%) | 15(50%) | 30 |
| Sex of the head | Male | 48(51.1%) | 46(48.9%) | 46(48.9%) | 48(51.1%) | 94 |
|  | Female | 6(19.4%) | 25(80.6%) | 6(19.4%) | 25(80.6%) | 31 |
| Educational status of the family head | Illiterate | 12(60%) | 8(40%) | 12(60%) | 8(40%) | 20 |
|  | Primary | 12(57.1%) | 9(42.9%) | 11(52.4%) | 10(47.6%) | 21 |
|  | Secondary | 8(33.3%) | 16(66.7%) | 8(33.3%) | 16(66.7%) | 24 |
|  | College& above | 22(36.7%) | 38(63.3%) | 21(35%) | 39(65%) | 60 |
| Occupation of the family head | Farmer | 11(61.1%) | 7(38.9%) | 11(61.1%) | 7(38.9%) | 18 |
|  | Gov't employer | 15(31.3%) | 33(68.8%) | 15(31.3%) | 33(68.8%) | 48 |
|  | Self-employer | 12(48%) | 13(52%) | 12(48%) | 13(52%) | 25 |
|  | Daily laborer | 5(62.5%) | 3(37.5%) | 4(50%) | 4(50%) | 8 |
|  | House wife | 2(25%) | 6(75%) | 2(25%) | 6(75%) | 8 |
|  | Merchant | 9(50%) | 9(50%) | 8(44.4%) | 10(55.6%) | 18 |
| Marital status of family head | Married | 42(47.7%) | 46(52.3%) | 41(46.6%) | 47(53.4%) | 88 |
|  | Single | 7(30.4%) | 16(69.6%) | 6(26.1%) | 17(73.9%) | 23 |
|  | Divorced | 4(33.3%) | 8(66.7%) | 4(33.3%) | 8(66.7%) | 12 |
|  | Widowed | 1(50%) | 1(50%) | 1(50%) | 1(50%) | 2 |
| Family income in birr | 500-1000 | 10(66.7%) | 5(33.3%) | 9(60%) | 6(40%) | 15 |
|  | 1001-3000 | 10(58.8%) | 7(41.2%) | 10(58.8%) | 7(41.2%) | 19 |
|  | 3001-5000 | 7(33.3%) | 14(66.7%) | 7(33.3%) | 14(66.7%) | 21 |
|  | >5000 | 27(37.5%) | 45(62.5%) | 26(36.1%) | 46(63.9%) | 70 |
|  | 1-3 | 26(40.6%) | 38(59.4%) | 25(39.1%) | 39(60.9%) | 64 |
| Family size | 4-5 | 14(40%) | 21(60%) | 14(40%) | 21(60%) | 35 |
|  | >5 | 14(53.8%) | 12(46.2%) | 13(50%) | 13(50%) | 26 |

### Factors associated with bacteriological quality of drinking water in Debre Markos town, Ethiopia

Table 5 indicated the presents of hand washing facilities near the toilet observed at the majority of households (69.6%), while 30.4% of households had not hand washing facilities. Most of respondents’ households (70.4%) were using soap/ash to wash hands after using toilet and 64.8% of respondents’ households had a waste container.

**Table 5:**
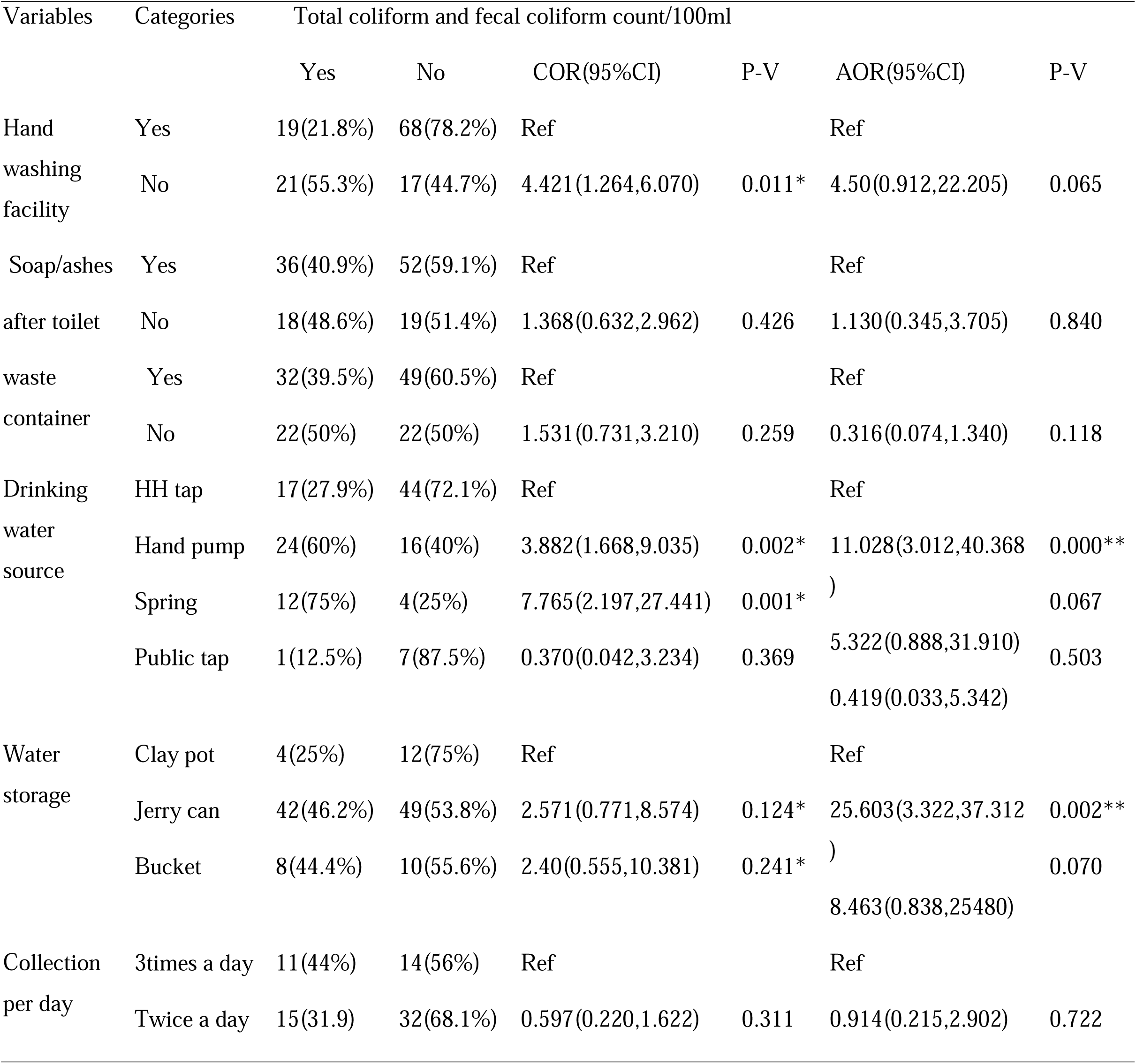
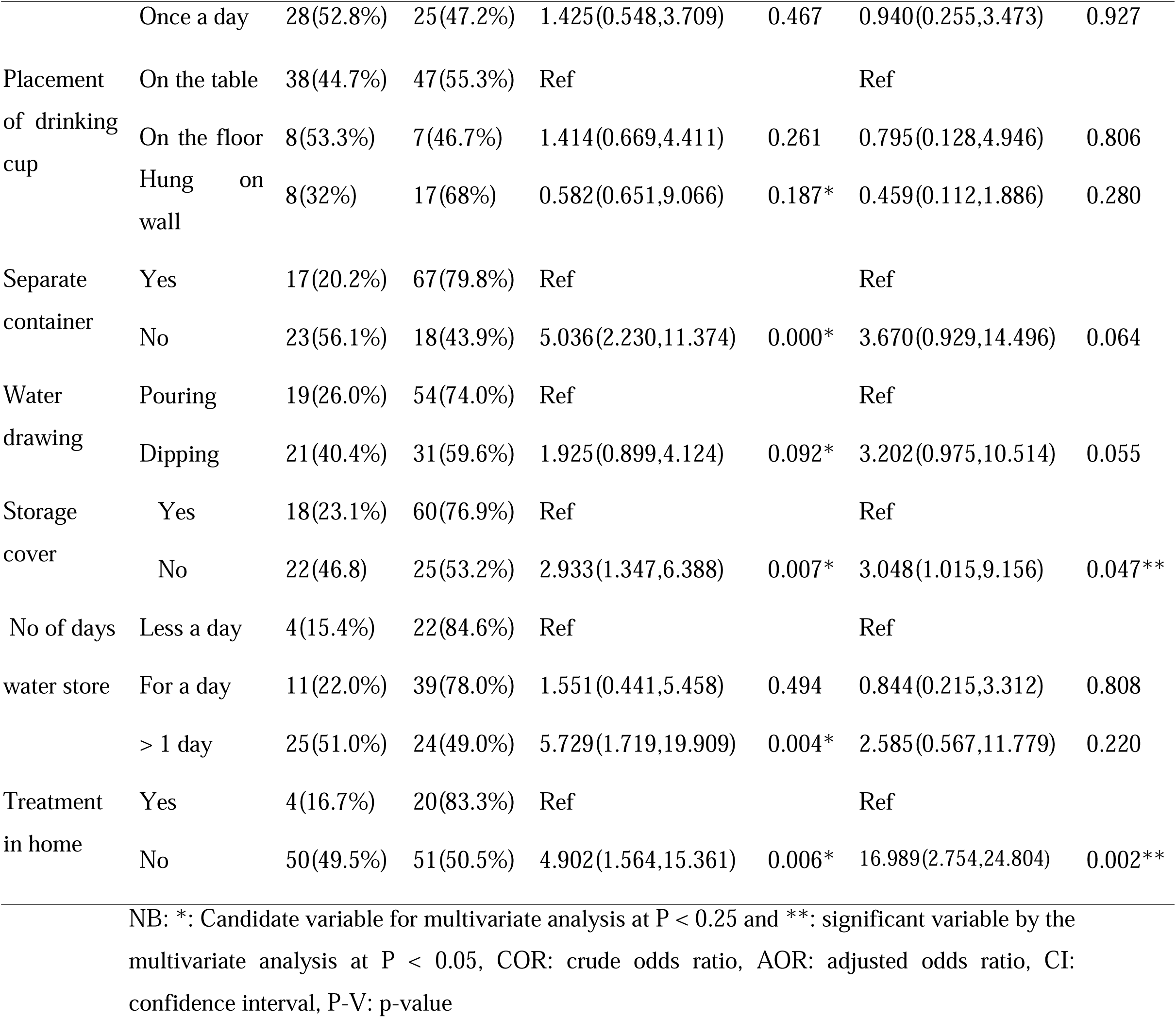
Bivariate and Multivariate logistic regression of associated factors for bacteriological quality of drinking water in Debre Markos town, Ethiopia.

Bivariate and multivariate analysis were done in logistic regression to detect factors associated with the bacteriological quality of drinking water. In bivariate logistic regression analysis, no hand washing facility near the toilet, hand pump and spring water source, water storage materials (jerry can and plastic bucket) were significantly associated with bacteriological quality of drinking water. In addition, no separate container for drinking, no storage cover, dipping method of water drawing, drinking cup hung on the wall, water store more than a day and no water treatment practice in home were significantly associated with bacteriological quality of drinking water.

Indeed, multivariate logistic regression analysis was also carry out after adjusting the potential confounding effects to identify significantly associated factors. In multivariate logistic regression, drinking water from tap water, wide container for water storage, storage containers have cover and usage of water treatment in home were protective factors against having contaminated drinking water at the point-of-use (Table 5).

## DISCUSSION

The microbiological examination of drinking water emphasizes on quality of water supply and assessment of hygiene [17]. This requires the detection of organisms that indicate the presence of fecal contamination. The WHO recommends that water used for human drinking purpose should be free from microbial contamination. The detection of thermotolerant/fecal coliform is a strong evidence for fecal contamination and a potential health risk for consumers, while a high level of total coliform at least implies inadequate chlorination that in turn could lead to bacterial contamination of the drinking water [1].

In this study, we found that 84/211(39.8%); (95% CI: (37-53)) and 75/211(35.5%); (95% CI: (31-47)) of all drinking water samples were positive for total coliforms and fecal coliforms respectively. This finding is in line with a study conducted in northern India that 42.9% of water samples were positive for TC and TTC [10]. This finding is higher than the results reported from Addis Ababa [18] 7% and 3% of water samples were positive for TC and TTC respectively and Bangladesh [19] 10.5% of samples were positive for TC and TTC. These variation may be due to difference in; geographical area, socioeconomic status, hygienic practices and drinking water handling practices. In other words, this finding was lower than the studies reported from Dire Dawa (83.3%) [20], Jimma (87.5%) [21] and Adis Kidame town [22] that 89% of drinking water were positive for TC and 77% of samples were positive for TTC. These variation may be due to difference in; geographical area, socioeconomic status, type of laboratory method used, sample size, sampling technique, hygienic practices and drinking water handling practices.

The bacteriological quality of drinking water is vary from source to source and from sources to point of use [23]. In the current study, water sources were contaminated by TC and TTC respectively with 13 (52%); (95% CI: (31-73)) and 6(24%); (95% CI: (6-42)) of the total drinking water sources. This study also revealed that 42%; (95% CI: (33-50)) and 43%; (95% CI: (34-52)) of household’s container were contaminated with TC and TTC respectively and 28%; (95% CI: (16-39)) of tap water were positive for both TC and TTC. This finding is not in compliance with the WHO and Ethiopian standards for drinking water quality (0 TTC/ 100ml water sample). The finding of TTC in present study from water sources is in line with previous studies reported from Kenya (17.5%) [24], Eastern Ethiopia (21.7%) [25] and Jimma (35.7%) [21]. This finding is lower than studies reported from Fogera (74%) [26], North Gondar Zone (56.5%) [16] and from other studies reported in Ethiopia (80 to 100%) [8, 27]. The finding of TC from sources in the present study is higher than the results reported from Eastern zone, Tigrai, Ethiopia (3.4%) [5] and Adama town, Ethiopia (5.8%) [28]. This discrepancy might be due to the difference of the season when the study was done, difference water sources, difference of water management and environmental sanitation.

In this study, boreholes were zero value of thermotolerant coliforms; in contrast, as the study conducted in Munsa woreda [29] indicated that high level of thermotolerant coliforms were recorded. This difference might be due to the lack of water sources protection in the case of Munsa woreda and not in case of Debre Markos town, the area is protected by wire fence. Reservoirs were zero value of TC and TTC, this might be due to the location of boreholes those are found very far from residential area and use of adequate chlorination on treatment plant. The presence of TC from boreholes might be due to naturally. On the other hand, public water sources except public taps, value of TC and TTC were above the recommendation limit of Ethiopian standard and WHO guideline. This might be due to the location of water sources those are found near to the residence and inadequate treatment. The zero value of TC and TTC of public taps might be due to its water supply network, the water supply is from the main sources of the town similarly to household tap and got adequate treatment in the reservoirs.

In the current study, the contamination of water by TC and TTC in household taps were similar, 17(28%); (95% CI: (16-39)). This finding is in line with studies done in Jijiga (26.7%) [30] and Nekemte (37%) [31]. A study in Bahir Dar [32] reported that 77.1% of tap water samples were positive for TC, which is higher than the current study and the discrepancy might be due to the difference of water source supply and season variation. This 28% of TC and TTC contamination in faucet (tap) in the present study, might be due to water pick up contaminants on its journey, while reservoirs were free from indicator bacteria. This may be likely due to the damage of the waterline and the possible entry of contaminants [33].

In present study, regarding the bacteriological quality of water source type, 30% of hand-pumps, and 50% of protected springs, 100% of unprotected springs and 28% of household tap water samples were contaminated by thermotolerant coliforms. Hand pumps and springs water sources were more contaminated than tap water. The variation of contamination between water sources might be due to its location, fence and treatment facilities. According to risk classification for thermotolerant coliforms88, mean value thermotolerant coliforms in the taps, household’s containers and springs were in the low-risk categories. However, mean value TTC from hand pumps were in high-risk categories. Regarding the boreholes, reservoir and public tap, water samples were at no-risk category. Generally based on the CFU/100ml, results did not comply with WHO guidelines [1]. From total sources 19 (76%) had <1 TTC/100 ml and 6 (24%) of the sources had ≥ 1 TTC/100 ml. Unprotected water sources were more contaminated than the protected water sources and the proportion of 1–10 to 11–50 CFU/100 ml water count is higher for protected water source than unprotected. This might be due to that protected water sources are not allowed the entrance of animal, continuously openly flowed and cleaned environment of protected water. Studies from Farta woreda [34] and Boloso Sore Woreda [8] also supported this finding.

This study revealed that 43%; 95% CI (34%, 52%) and 42%; 95% CI (33%, 50%) of households drinking water were contaminated with TC and TTC, respectively. This finding is in line with study reported from Adama (42.3%) [28] and from Jijiga (41.7%) [30] of households’ water were contaminated with TTC. This finding is lower than the result reported from Ghana (83%) [35] Eastern Ethiopia (83.3%)60 North Gonder zone (72.6%) [36] and Farta woreda (100%) [34]. This variation might be explained by difference in; geographical area, type of drinking water sources, water handling practices in related to collection, and transportation and hygienic practices related to storage, and usage of water in household.

In the current study, associated factors were also assessed and there are some independent variables associated with bacteriological quality of drinking water. Among these predictors, source of water, water storage material, storage cover and water treatment in home were identified as important determinants for bacteriological quality of drinking water. Households those benefited drinking water from hand pump had more contaminated water samples by indicator bacteria than those of households benefited from household tap (AOR=11; 95% CI: (3.012-40.368)) (p<0.001). This finding showed that respondents that benefited drinking water from hand pump was 11 times more likely to be contaminated than those households that benefited from household tap (AOR=11; 95% CI: (3.012-40.368)) (p<0.001). This finding was harmony with the finding in Eastern zone, Tigray, Ethiopia [5] that revealed water samples of hand pump were contaminated by indicator bacteria and associated with inadequate fencing around the sources, however, in present study the association might be due to inadequate treatment of sources.

Study participants were used jerry can (72.8%), clay pot (12.8%) and plastic bucket (14.4%) to store drinking water. The majority of the respondents use Jerry can for both the collection (78.4%) and storage (72.8%). Households used not covered jerry can to store water were 25.6 times more contaminated than households used covered clay pot (AOR=25.6; 95% CI: (3.322-37.312)) (p<0.002). This might be due to jerry can have a very narrow space through which enough oxygen cannot get in [37]. This study showed that drinking water from households that have no storage cover was 3 times more likely to be contaminated than those households that have storage cover (AOR=3; 95% CI: (1.015-9.156)) (p<0.047). The main contribution for household water contamination were related to storage and usage activities such as: selection of household containers, not covered water storage container and not used household water treatment, because of these factors the thermotolerant coliform load increases by two third folds in household containers than sources [38].

This study showed that drinking water from households that did not practice household water treatment in home was 16.9 times more likely to be contaminated than those households that practice household water treatment in home (AOR=16.9; 95% CI: (2.754-24.804)) (p<0.002). This could be explained by the fact that treating water at home, use will help to improve the quality of drinking water by attacking the microorganisms and the risk of contamination could be prevented. In this study, 80.8% of the respondents did not treat drinking water in home, this was in accordance with the study done in rural community of Dire Dawa [39] where 87% of the respondents did not treat their water at household level.

In generally, drinking water at the households (42%) were more contaminated with TTC compared to that of the sources (24%). This finding is agreed with study done in Kobo town [40] that showed water storage and usage containers affected the household water quality that showed 100% of households and 93.3% of sources were contaminated with TTC. Research done in Boloso Sore woreda [8] also showed that households (91%) was more contaminated than sources (44%) with TTC. In addition, drinking water from household containers were found more contaminated by coliforms compared to drinking water from tap waters. This might be due to that water can be contaminated with coliform both in pipe during journey and in households. This occur in households those used narrow container for storage rather than wide storage container, if storage container were not covered and if households were not treat drinking water in home [32].

## CONCLUSION

In conclusion, bacteriological quality of drinking water in Debre Markos town fails to comply with WHO guidelines and national standards. This study result indicates that bacteriological quality of drinking water deteriorates from sources to point of use. The quality was declined after collection from sources. Furthermore, type of drinking water source, selection of household water storage container, storage cover and usage of household water treatment were associated factors of bacteriological quality of drinking water among households. The presence of total coliform in drinking water indicate inadequate chlorination and the presence of thermotolerant coliforms in the drinking water samples demonstrates the presence of pathogenic microorganisms that would be a threat to individual consuming the water.

## Data Availability

All data produced in the present study are available upon reasonable request to the authors

https://drive.google.com/file/d/1RL8HyWOweoSUcQ8Yi2kmV9dQthodH-at/view?usp=drive_link

## ETHICS APPROVAL

This study was ethically approved by the research and ethical review committee of College of Health Science, Debre Markos University with reference number DMU/CHS/RERC/278/13. It was performed in accordance with the relevant guidelines and regulations.

## CONSENT TO PARTICIPATE

Before sample collection and written consent was obtained from each study participants.

## AVAILABILITY OF DATA

The datasets used and/or analyzed during the current study are available from the corresponding author on reasonable request.

## ACKNOWLEDGMENTS

We acknowledge Debre Markos town water supply and sewerage service and study participants.

